# Brainstem Auditory Physiology in Children with Listening Difficulties

**DOI:** 10.1101/2022.07.12.22277544

**Authors:** Lisa L. Hunter, Chelsea M. Blankenship, Barbara Shinn-Cunningham, Linda Hood, Lina Motlagh Zadeh, David R. Moore

**Affiliations:** Communication Sciences Research Center, Cincinnati Children’s Hospital Medical Center, Cincinnati, Ohio. USA; Research in Patient Services, Cincinnati Children’s Hospital Medical Center, Cincinnati, Ohio. USA; College of Medicine, Otolaryngology, University of Cincinnati, Cincinnati, Ohio. USA; College of Allied Health Sciences, Communication Sciences and Disorders, University of Cincinnati, Cincinnati, Ohio. USA; Neuroscience Institute, Carnegie Mellon University, Pittsburgh, PA. USA; Vanderbilt University Medical Center, Nashville, TN. USA; Manchester Centre for Audiology and Deafness, University of Manchester, U.K.

**Keywords:** Listening difficulty, auditory processing disorder, middle ear muscle reflex, auditory brainstem response, speech in noise, extended high frequency.

## Abstract

Children who have listening difficulties (LiD) despite having normal audiometry are often diagnosed as having an auditory processing disorder (APD). A lack of evidence regarding involvement of specific auditory mechanisms has limited development of effective treatments for these children. Here, we examined electrophysiologic evidence for brainstem pathway mechanisms in children with and without defined LiD. We undertook a prospective controlled study of 132 children aged 6-14 years with normal pure tone audiometry, grouped into LiD (n=63) or Typically Developing (TD; n=69) based on scores on the Evaluation of Children’s Listening and Processing Skills (ECLiPS), a validated caregiver report. The groups were matched on age at test, sex, race, and ethnicity. Neither group had diagnoses of major neurologic disorder, intellectual disability, or brain injuries. Both groups received a test battery designed to measure receptive speech perception against distractor speech, Listening in Spatialized Noise - Sentences (LiSN-S), along with multiple neurophysiologic measures that tap afferent and efferent auditory subcortical pathways. Group analysis showed that participants with LiD performed significantly more poorly on all subtests of the LiSN-S. The LiD group had significantly steeper wideband middle ear muscle reflex (MEMR) growth functions and shorter Wave III, Wave V, and I-V interpeak latencies in their auditory brainstem responses (ABR). Across individual participants, shorter latency ABR Wave V correlated significantly with poorer parent report of LiD (ECLiPS composite). Steeper MEMR growth functions also correlated with poorer ECLiPS scores and reduced LiSN-S talker advantage. The LiD and TD groups had equivalent summating potentials, compound action potentials, envelope-following responses (EFR), and binaurally activated medial olivocochlear reflexes (MOCR). In conclusion, there was no evidence for auditory synaptopathy or lower brainstem dysfunction for LiD. Evidence for higher brainstem differences between groups showed that the LiD group had increased efferent control or central gain, with shorter ABR Wave III and V latencies and steeper MEMR growth curves. These differences were related to poorer parent report and speech perception ability.

## 1.0 INTRODUCTION

Children with unexplained listening difficulties (LiD) that interfere with learning despite having clinically normal peripheral hearing, may be diagnosed as having auditory processing disorder (APD), a condition that often co-exists with developmental language disorder and attention deficit hyperactivity disorder (Dawes & Bishop, 2009; Ferguson, Hall, Riley, & Moore, 2011; Miller & Wagstaff, 2011; Moore, Ferguson, Edmondson-Jones, Ratib, & Riley, 2010; Sharma, Purdy, & Kelly, 2009). For children with LiD defined by validated caregiver report, we recently found that the peripheral auditory system was functioning normally, except for a subgroup with poorer extended high frequency (EHF) hearing (Hunter, Blankenship, et al., 2020b), while supramodal cognitive factors (e.g. language, memory and attention) were highly correlated with their listening difficulties (Petley et al., 2021).

Neurophysiologic techniques have the advantage of identifying underlying mechanisms without relying upon behavioral responses. They have thus been recommended to evaluate auditory pathway involvement in APD (AAA, 2010; Ankmnal-Veeranna, Allan, & Allen, 2019; ASHA, 2005; Jerger & Musiek, 2000). However, studies using these techniques have produced inconsistent findings, probably due to varying definitions of APD, retrospective design, inadequate methodologic control, small group sizes, or lack of a control group (Liu, Zhu, Chen, Hong, & Chi, 2021).

Ascending auditory neuronal transmission can be investigated from the inner hair cells to the lateral lemniscus using electrocochleographic (ECoG) and auditory brainstem responses (ABR). Temporal encoding has been investigated using narrow- (Bharadwaj, Verhulst, Shaheen, Liberman, & Shinn-Cunningham, 2014) and multi-band (Wang, Bharadwaj, & Shinn-Cunningham, 2019) complex tones to measure envelope following responses (EFR) driven by specific frequency regions of the cochlea. These responses have been proposed as a neurophysiological measure for detecting subtle temporal coding deficits in listeners with normal hearing who have difficulty understanding target sounds in complex acoustic scenes, such as might arise with cochlear synaptopathy (Encina-Llamas, Harte, Dau, Shinn-Cunningham, & Epp, 2019; Shaheen, Valero, & Liberman, 2015). Deficits in spectrotemporal processing have been reported in children with APD and are associated with poorer speech in noise perception (Ankmnal Veeranna, Allan, Macpherson, & Allen, 2019; Lotfi, Moossavi, Afshari, Bakhshi, & Sadjedi, 2020), although such deficits also present in visual temporal processing, so may reflect general processing speed problems (Dawes et al., 2009). Thus far, EFR has not been investigated in children with LiD and could shed light on neural mechanisms of auditory temporal processing in children with LiD.

Multimodal pathways, deriving from anterior temporal, and inferior frontal and parietal cortex interact with ascending processing in the auditory cortex, giving rise to a large descending auditory system (Hackett, 2011; Moore, Rosen, Bamiou, Campbell, & Sirimanna, 2013). One important function of such cortical and sub-cortical efferent pathways is to modulate ascending sensory auditory information from the ear. Recent evidence from animal studies suggests that top-down pathways to subcortical auditory nuclei (medial geniculate and inferior colliculus) are influential in challenging listening situations (Blackwell, Lesicko, Rao, De Biasi, & Geffen, 2020; Souffi, Nodal, Bajo, & Edeline, 2021). Our striking human ability to maintain speech understanding in highly degraded listening conditions of noise, reverberation, and competing meaningful speech (the “cocktail party” effect) depends in part on these efferent cortical pathways, which modulate neural activity in the inferior colliculus (Hernandez-Perez et al., 2021).

The auditory efferent system can be assessed with the medial olivocochlear reflex (MOCR) and middle ear muscle reflexes (MEMR). The MEMR pathway begins in the auditory periphery (middle ear cochlea, auditory nerve) and projects to the cochlear nucleus in the brainstem, the first ascending central auditory relay station. From there, ventral cochlear nucleus interneurons activate efferent stapedius motoneurons and initiate a muscle contraction in the same ear (ipsilateral) and, also in the contralateral ear, as the reflex is consensual, like the pupillary reflex (Mukerji, Windsor, & Lee, 2010). Thus, the MEMR includes both ascending afferent pathways and descending efferent pathways to the same and opposite ears. Some children with LiD exhibit shallower MEMR growth as a function of stimulus intensity (Saxena, Allan, & Allen, 2015), but MEMR thresholds were no different in children diagnosed with APD compared to those without this diagnosis (Allen & Allan, 2014). We recently found that children with LiD and a control group showed no differences in MEMR thresholds using a wideband absorbance technique (Hunter, Blankenship, et al., 2020a). In contrast to shallower growth, patients exposed to aminoglycoside antibiotics recently were shown to have greater MEMR growth than non-exposed controls, despite having normal pure tone thresholds (Westman, Putterman, Garinis, Hunter, & Feeney, 2021). The steeper growth was interpreted as evidence for central gain, consistent with animal and temporal bone studies showing synaptic damage with aminoglycoside exposure.

MOCR strength has been quantified using transient-evoked otoacoustic emissions (TEOAEs) in quiet and with monaural and binaural elicitors in a forward masking paradigm (Berlin, Hood, Hurley, Wen, & Kemp, 1995). MOCR dysfunction has been suggested to affect speech perception due to longer cochlear ringing and increases in forward masking (Boothalingam, Allan, Allen, & Purcell, 2015). The MOCR may be selectively activated by distorted (e.g. vocoded) speech rather than by clear speech in noise (Hernandez-Perez et al., 2021). Several studies have examined MOCR in children with suspected APD or LiD, but results have been conflicting. An evidence-based review of nine such studies in children with APD, dyslexia, learning impairment, or specific language impairment showed that the MOCR was reduced in DPOAE or TEOAE suppression relative to controls in 4 studies, while 3 studies showed no significant group differences (Mishra, 2014). Variability across studies is likely due to subject selection, methodological differences and test-retest variability (Boothalingam, Allan, Allen, & Purcell, 2019; Mertes & Goodman, 2016; Mertes & Leek, 2016), inadequate control of OAE signal-to-noise ratio (SNR), attention effects (Mishra, 2014), or unintended activation of the MEMR (Marks & Siegel, 2017; Mishra, 2014).

To examine mechanisms that may underly LiD, we conducted a prospective evaluation of afferent and efferent auditory brainstem function in children with and without defined LiD using neurophysiologic tests selected to investigate specific regions of the auditory pathway. We hypothesized that children with LiD have deficiencies in either ascending or descending auditory brainstem pathways that relate to their speech-in-noise deficits.

## 2.0 MATERIALS AND METHODS

### 2.1 Participants

The study was approved by the Cincinnati Children’s Hospital (CCH) Institutional Review Board. Parental permission and child assent for those 11 years and older was obtained before assessments. This report is part of a broader longitudinal Sensitive Indices of Childhood Listening Difficulty (SICLiD) study that aimed to uncover mechanisms of LiD; enrollment methods were thoroughly described previously in Petley et al., 2021a. Briefly, children with LiD aged 6 to 14 years old at enrollment were age- and gender-matched to typically developing (TD) children by proportional sampling. The LiD group was recruited from clinical services and website advertisements at CCH, and enrolled based on low (< -1 s.d.) age-standardized total scores on the Evaluation of Children’s Listening and Processing Skills (ECLiPS), a validated parent questionnaire (Barry & Moore, 2021; Barry, Tomlin, Moore, & Dillon, 2015). Inclusion criteria for both groups were no major neurologic or cognitive diagnoses, no brain injuries, normal bilateral standard frequency pure tone hearing thresholds (≤20 dB HL; .25-8 kHz), normal otoscopy, and normal 226-Hz tympanometry (tympanometric width < 250 daPa).

Study data were collected and managed using Research Electronic Data Capture (REDCap) electronic data capture tools hosted at the University of Cincinnati (Harris et al., 2019; Harris et al., 2009).

### 2.2 Procedures

#### Audiological Assessment

Otoscopy was completed and if necessary, cerumen was removed before audiometry. All audiometric tests were completed in a double-walled soundproof booth (Industrial Acoustics Company, North Aurora, IL) that meets standards for acceptable room noise for audiometric rooms (ANSI/ASA 1999 (R2018). Standard (0.25 to 8 kHz) and EHF (10–16 kHz) thresholds were measured using the manual Hughson–Westlake method for the range of 0.25 to 8 kHz at octave intervals and at four additional frequencies (10, 12.5, 14, and 16 kHz) using an Equinox audiometer (Interacoustics Inc., Middlefart, Denmark) with Sennheiser HDA-300 circumaural earphones (Old Lyme, CT). Calibration was completed according to ISO 389.9 (International Organization for Standardization, 2004) for standard frequencies and ISO 389-1 (International Organization for Standardization, 2017) for EHF. Normal hearing was defined as thresholds ≤ 20 dB HL (0.25-8 kHz). If air conduction thresholds were greater than 20 dB HL, bone conduction thresholds were measured between 0.5 and 4 kHz using appropriate narrowband masking in the contralateral ear (Radioear Inc. B-71 bone vibrator, New Eagle, PA) to determine the type of hearing loss.

#### Speech in Noise

The Listening in Spatialized Noise – Sentences (LiSN-S) task, North American version (Brown, Cameron, Martin, Watson, & Dillon, 2010; Cameron & Dillon, 2007) was administered using a laptop, a task-specific soundcard, and Sennheiser HD 215 headphones. Participants were asked to repeat a series of target sentences, presented from directly in front (0°), while ignoring two distracting sentences. There are four listening conditions, in which the distractors change voice (either different or the same as target) and/or position (either both at 0° or at -90° and +90° degrees relative to the listener). The test is adaptive; the level of the target speaker decreases or increases in SNR depending on listener accuracy. Each condition continues for 22-30 trials, ending when the standard error of reversals is < 1 dB. The 50% correct SNR is calculated for the ‘Low cue speech reception threshold’ (SRT; same voice, 0° relative to the listener) and the ‘High cue SRT’ (different voice, 90° degrees relative to the listener). Three ‘derived scores’ are the Talker Advantage (difference in thresholds for different voice vs. same voice when the distractors are at 0°), Spatial Advantage (difference in thresholds for spatially separated vs. spatially collocated distractors when the voice is the same), and Total Advantage (difference in the High cue SRT and the Low cue SRT).

#### Middle Ear Muscle Reflex

Wideband tympanometry was performed prior to MEMRs (acoustic absorbance and group delay) using click stimuli (bandwidth 0.2 to 8 kHz) delivered while ear canal pressure was swept from +200 to −400 daPa using a custom recording system (Keefe, Feeney, Hunter, & Fitzpatrick, 2017) coupled to an AT235 immittance system (Interacoustics Inc., Middlefart, Denmark). MEMRs were measured using the wideband absorbance technique with custom MATLAB software described by Keefe et al. (2017). The probe assembly contained a high bandwidth receiver that delivered wideband clicks as the probe stimulus, and a second receiver with the same bandwidth that allowed higher stimulus levels. Broadband noise (BBN, 0.2 to 8 kHz) and pure-tone stimuli (0.5, 1, and 2kHz) were presented ipsilaterally and contralaterally to the probe ear while the click stimulus was presented ipsilaterally to measure absorbance changes in the ear with the microphone. Ear canal air pressure was adjusted to the average peak tympanometric pressure obtained for down swept and upswept wideband tympanometry. To record MEMR responses, probe clicks were averaged across four stimuli, calibrated in a 2-cc coupler and in the real ear. Contralateral and ipsilateral MEMR testing used response averaging, artifact rejection and signal processing techniques to measure threshold, onset latency, and amplitude growth with click level. Amplitude growth of the MEMR was recorded at 10 levels (L1-L10, where L1 is the lowest level). For BBN, MEMRs were measured at 0 dB SPL and then from 50 to 90 dB SPL in 5 dB steps. Similarly, for pure-tones, MEMRs were measured at 0 dB SPL and then 65 to 105 dB SPL in 5 dB steps. At each stimulus level, the MEMR shift was measured in cumulative weighted absorbed power level in dB, averaged across all frequencies (0.2 to 8 kHz). Next, this value was used to calculate the MEMR Slope (L10-L2/40).

#### Electrocochleography and Auditory Brainstem Response

Combined ECoG and ABR recordings were obtained using the Intelligent Hearing Systems (IHS, Miami, FL) SmartEP two-channel system with the universal SmartBox platform. IHS ultra-shielded insert earphones (300Ω) coupled to gold leaf tiptrodes were used to deliver stimuli and simultaneously to serve as the negative electrodes. All recordings were performed in a double-walled sound booth. Stimuli were alternating clicks, split into rarefaction and condensation buffers at 75, 80, 85 and 90 dB nHL, at a rate of 11.1 clicks/sec, with 2048 sweeps recorded per intensity, and repeated for a total of 4096 sweeps per intensity. Filters were 0.1-3 kHz with gain of 100k. The two-channel recording montage was high forehead (positive) to bilateral ear canals, with the ground at the low forehead. Care was taken to insert the tiptrodes as deeply as possible without disturbing the gold foil. Impedances were maintained at less than 5 kOhms, and <2 kOhms difference between electrodes. If peaks were not easily discernable or the number of artifacts was greater than or equal to the 10% of the number of sweeps, the recording was repeated, and the best two recordings (best defined peaks with least noise) were analyzed. Ipsilateral and contralateral recordings were analyzed using a normative latency/intensity template to guide selection for consistent marking of latency and amplitude of the summating potential (SP; base to shoulder), and Waves I, III and V (peak to the negative trough following the waveform). Recordings were manually marked with blinding of subject information by one investigator and were independently cross-checked by a blinded second scorer to ensure agreement on marked latencies and amplitudes. Any discrepancies were reviewed by the first author (LLH) with blinding maintained. Sixteen participants were excluded due to inadequate ABR quality (8 LiD, 8 TD).

#### Binaural Medial Olivocochlear Reflex

The MOCR was measured using TEOAEs and a binaural elicitor with an Intelligent Hearing Systems dual channel universal SmartBox (SmartTrOAE, Miami, FL) with two matched OAE 10D Probes. All recordings were performed in a double-walled sound booth. TEOAEs were first recorded in quiet with non-linear rectangular clicks (three clicks of positive polarity followed by a fourth click of inverse polarity with a relative magnitude of 9.5 dB higher than the corresponding positive clicks) in each ear (75 µsec, 80 dB peSPL, 21.1 per sec). To ensure that baseline responses were present, 3 dB SNR for 3 or more frequency bands out of 6, and with >60% correlation, or whole wave reproducibility was required, and artifacts were required to be below 10% of the total. The MOCR was then elicited by recording TEOAEs with clicks (75 µsec, 60 dB peSPL, 21.1 per sec) with 256 sweeps in each condition. The 60 dB peSPL TEOAE activator and elicitor levels were set to be below MEMR thresholds to minimize activating the stapedial reflex during MOCR measurement. The activator and elicitor stimuli were interleaved with order of testing as follows: Quiet Condition 1; Binaural Elicitor Condition 1; Quiet Condition 2; Binaural Elicitor Condition 2. The elicitor was binaural white noise at 60 dB SPL, presented in a forward masking paradigm, with a 400 ms. duration elicitor, and an interstimulus interval of 10 ms. If the SNR was <3 dB or the correlation was < .6, then an additional 256 sweeps were recorded (512 total). Artifacts were required to be less than 10% of the total. A reclining chair was used to encourage participants to be still and quiet. Attention was controlled by having the subject watch a silent video with captions. If the recording did not meet SNR criteria, the probe was refit, and the participant was reminded to remain quiet and still. Recordings were analyzed using the IHS MOCR analysis module set to a 10 ms. window from 8 to 18 ms., Hanning filter, 2 ms. resolution, and coherence display setting. The RMS amplitude for each waveform was recorded, and the with elicitor condition was subtracted from the without elicitor (quiet) condition to obtain the average MOCR strength estimate for each ear.

#### Envelope Following Response

The EFR was recorded using a multi-channel actiCHamp Brain Products system (Brain Products GmbH, Inc., Munich, Germany). A 64-electrode cap was placed on the scalp with electrodes placed at equidistant locations. This “infracerebral” cap covers a larger area than is typical in a 10–20 system (Hine & Debener, 2007). The reference channel was located at vertex (Cz) while the ground electrode was located on the midline, 50% of the distance to nasion. Responses were recorded using a sampling rate of 5 kHz. The stimuli were transposed tones generated offline in MATLAB (Natick, MA) and stored for playback with a sampling rate of 48.8 kHz. Stimuli were presented at three different modulation depths, 100%, 63% and 40%, with a modulation rate of 100 Hz and carrier frequency of 4 kHz (Bharadwaj et al., 2014). The stimuli were 400 ms. in duration and 1000 trials of each modulation depth were recorded. The inter-trial interval was jittered between 410 and 510 ms. to ensure that EEG noise (not in response to the stimulus) occurs at a random phase between −π and π for frequencies above 10 Hz. Stimuli were presented diotically over ER-2 insert earphones (Etymotic Research, Elk Grove Village, IL) with the level at each ear at 70 dB SPL. All recordings were performed in a Faraday shielded double-walled sound booth. Participants were encouraged to sleep during testing, which took approximately 45 minutes to complete.

Electrophysiological data were analyzed using Brain Vision Analyzer ver. 2.0 (Brain Products GmbH, Inc., Munich, Germany) and custom Python and MATLAB scripts. To minimize signal contributions from cortical sources before epoching and to remove 60 Hz line noise, data were re-referenced to the left and right average mastoid reference and high-pass filtered in MATLAB with a 70 Hz cutoff frequency using an FIR filter with zero group-delay (Herdman et al., 2002; Kuwada et al., 2002). Response epochs from −50 to 250 ms (relative to the stimulus onset time of each trial) were segmented out from each channel. Epochs with signals whose dynamic range exceeded 100 μV in any channel were excluded from further analysis to remove movement and muscle activity artifacts. Principle component analysis was used to combine channels and reduce recording time. See Bharadwaj and Shinn-Cunningham (2014) for further details.

#### Statistical Analysis

For all measures, each recording was monitored online for excessive artifacts and noise and was repeated if necessary, during the same session, after taking care to obtain the quietest condition and best probe fit and/or electrode connection possible. Data were exported for each test, then were further analyzed for recording artifacts. If the test was repeated, the best quality recordings (lowest noise and artifacts) were selected for further analysis.

Statistical analysis was completed using JASP version 0.13.1 (University of Amsterdam). Results were examined initially with descriptive statistics to summarize sample demographics and outcome measurements. Interval variables were summarized by central tendency and dispersion, and categorical variables were described by frequencies and percentages. Two-sample t-tests and Chi-Square tests were used to compare the demographics between the children with LiD and TD. Boxplots were created to study the distribution of the outcomes. Outcome variables were analyzed first in univariate, then multivariate mixed models that included group (TD or LiD), age at test, sex, PE tube history, and EHF hearing loss (EHFHL) as independent factors. A two-sided significance level was set at p <0.05. Significant factors from the univariate analysis and between group demographics were included in the final multiple adjusted models, including all significant interaction effects. Holm multiple adjustment was applied for pairwise comparisons among the levels of the significant factors to maintain the experiment-wise error rate below alpha = .05 (Staffa & Zurakowski, 2020). Multivariate forward stepwise linear regressions were calculated to explore the relationship among the electrophysiologic and behavioral outcomes. P value <0.05 was required for entry to the model, and p >0.10 for removal.

## 3.0 RESULTS

### 3.1 Demographics

The current report includes 132 participants from the first SICLiD longitudinal evaluation (Table 1). There were no significant group differences in age at test, sex, race, ethnicity, or history of pressure equalization (PE) tube insertion to treat otitis media. The LiD group had significantly lower maternal education levels, more EHF hearing loss, and poorer scores on the ECLiPS (by selection) and LiSN-S, compared to the TD group.

**Table 1.** Study demographics for all participants, the typically developing (TD) group, and the listening difficulty group (LiD) group.

|  | All | TD | LiD | *p*-value |
| --- | --- | --- | --- | --- |
| **Number of Participants <sup>a</sup>** | 132 | 69 | 63 | — |
| **Age (yrs.) <sup>a</sup>** |  |  |  | 0.599 <sup>d</sup> |
| Mean (SD) | 10.0 (2.1) | 9.9 (2.1) | 10.1 (2.1) |  |
| Range | 6.2-14.6 | 6.3-14.6 | 6.2-14.1 |  |
| **Sex <sup>a</sup>** |  |  |  | 0.631 <sup>e</sup> |
| Male | 81 (61.4%) | 41 (59.4%) | 40 (63.5%) |  |
| Female | 51 (38.6%) | 28 (40.6%) | 23 (36.5%) |  |
| **Race <sup>a</sup>** |  |  |  | 0.172 <sup>e</sup> |
| Caucasian | 107 (81.1%) | 59 (85.5%) | 48 (76.2%) |  |
| Non-Caucasian | 25 (18.9%) | 10 (14.5%) | 15 (23.8%) |  |
| **Ethnicity <sup>a</sup>** |  |  |  | 0.550 <sup>e</sup> |
| Hispanic or Latino | 5 (3.8%) | 2 (2.9%) | 3 (4.8%) |  |
| Non-Hispanic or Latino | 125 (94.7%) | 67 (97.1%) | 58 (92.0%) |  |
| Prefer Not to Answer | 2 (1.5%) | 0 (0%) | 2 (3.2%) |  |
| **Maternal Education Level <sup>a</sup>** |  |  |  | ***0.001*** <sup>e</sup> |
| Graduated high school or less | 12 (9.1%) | 1 (1.4%) | 11 (17.5%) |  |
| Some college or more | 120 (90.9%) | 68 (98.6%) | 52 (82.5%) |  |
| **PE Tubes <sup>a</sup>** |  |  |  | 0.605 <sup>e</sup> |
| No History of Tubes | 100 (75.8%) | 51 (73.9%) | 49 (77.8%) |  |
| History of Tubes | 32 (24.2%) | 18 (26.1%) | 14 (22.2%) |  |
| **EHF Hearing Status <sup>b</sup>** |  |  |  | ***0.033*** <sup>e</sup> |
| Normal | 184 (74.5%) | 96 (73.8%) | 88 (75.2%) |  |
| Hearing Loss | 39 (15.8%) | 13 (10.0%) | 26 (22.2%) |  |
| Not Measured | 24 (9.7%) | 21 (16.2%) | 3 (2.6%) |  |
| **ECLIPS <sup>c</sup>** |  |  |  |  |
| Scaled Score | 7.1 (4.5) | 10.8 (2.5) | 2.9 (1.7) | ***<0.001*** <sup>d</sup> |
| **LiSN-S <sup>c</sup>** |  |  |  |  |
| Low-Cue | -0.5 (1.7) | -0.1 (1.0) | -1.0 (2.1) | ***<0.001*** <sup>d</sup> |
| High-Cue | 0.01 (1.2) | 0.4 (1.0) | -0.4 (2.1) | ***<0.001*** <sup>d</sup> |
| Talker Advantage | -0.3 (0.9) | -0.04 (0.8) | -0.5 (1.0) | ***<0.001*** <sup>d</sup> |
| Spatial Advantage | -0.3 (1.5) | -0.1 (1.2) | -0.6 (1.7) | ***0.002*** <sup>d</sup> |
| Total Advantage | 0.3 (1.2) | 0.5 (1.0) | 0.1 (1.3) | ***0.002*** <sup>d</sup> |

*Note:* <sup>a</sup> = number (%) of participants; <sup>b</sup> = number (%) of ears; <sup>c</sup> = mean (SD); <sup>d</sup> = Two-Sample t-test; <sup>e</sup> = Chi-Square test; Bold italics indicate significant *p*-values.

### 3.2 Electrocochleography and Auditory Brainstem Response

The ABR was selected as it is a highly reliable and diagnostic neurophysiologic measure, with low intrasubject variability when controlled for sex, as females generally have shorter latencies than males (McClelland & McCrea, 1979). Developmental trajectories reflect myelination and provide an indirect measure of brainstem travel time (Hecox & Burkard, 1982; Salamy, 1984). The difference between Wave V and Wave I is thought to reflect central conduction time, and has been found to be extended in children with autism (Miron et al., 2021; Rosenhall, Nordin, Brantberg, & Gillberg, 2003; Thivierge, Bedard, Cote, & Maziade, 1990; Wong & Wong, 1991).

There were significant decreases in ABR latency with increasing stimulus level for all waveform components, as expected (Figure 1 and Table 2). There was a significant right-left ear difference for SP latency (p=0.046), but no significant effects were found for group, age, EHF hearing level, or PE tube history on latency or amplitude, and there were no significant effects on Wave I (compound action potential). Thus, peripheral waveforms generated by the cochlea and auditory nerve were similar between groups. The LiD group had significantly shorter ipsilateral Wave III (p=0.001) and Wave V (p=0.036) ABR latencies than the TD group (Figure 1; C, D, G, H; Table 2). No other group differences for ipsilateral latency (Figure 1; A, B, E, F) or amplitude (Figure 2) were found. Group comparison of interpeak latencies for Wave I-III, III-V, and I-V; Figure 3) showed overall significantly shorter latencies for the LiD group (p = 0.012), accounted for by a shorter I-III travel time. As there was no difference in latency for Wave I, the shorter I-III interpeak latency indicates differences in travel time between the auditory nerve and the cochlear nucleus. The ipsilateral Wave V to Wave I amplitude ratio was not significantly different between groups (Table 2). There were no significant group differences in contralateral Wave V latency or amplitude (Figure 4).

**Figure 1.**
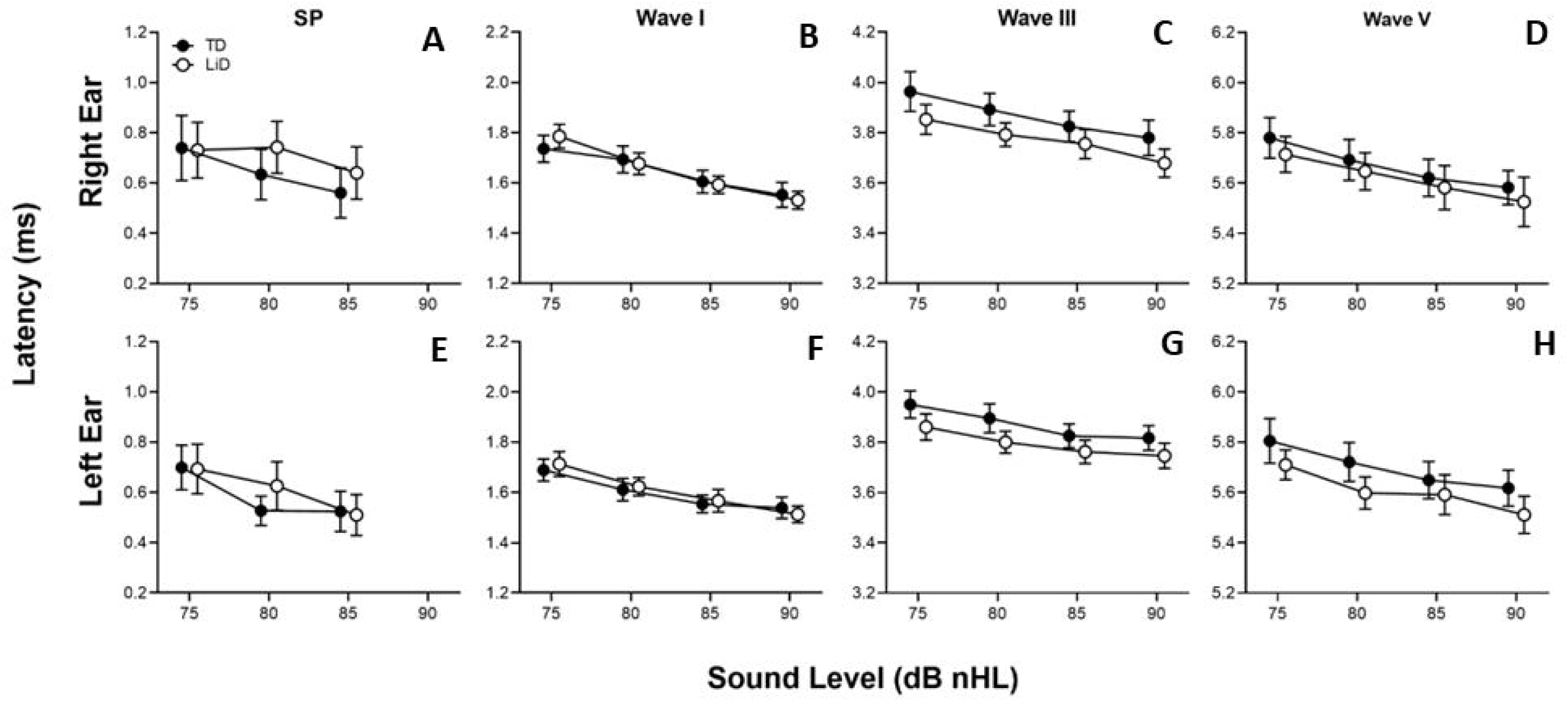
Ipsilateral ABR latency-intensity functions for the summating potential (SP), Waves I, III and V in dB HL re: normal adult threshold for clicks. The LiD and TD group are shown in open and filled circles, respectively. Error bars are 95% confidence intervals. N= 67 ears for the TD group and 66 ears for the LiD group.

**Figure 2.**
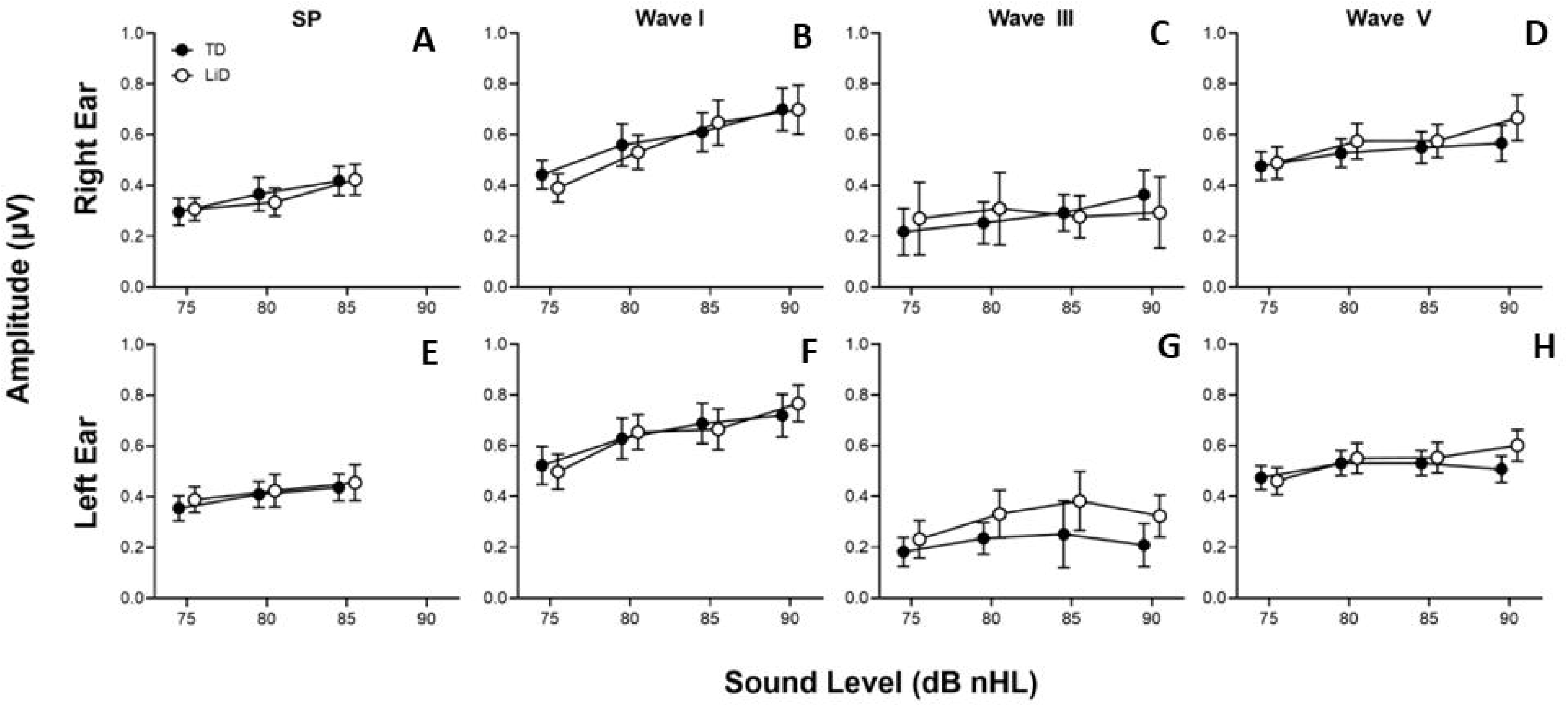
Ipsilateral ABR amplitude-intensity functions for the summating potential (SP), Waves I, III and V in dB HL re: normal adult threshold for clicks. The LiD and TD group are shown in open and filled circles, respectively. Error bars are 95% confidence intervals. N= 67 ears for the TD group and 66 ears for the LiD group.

**Figure 3.**
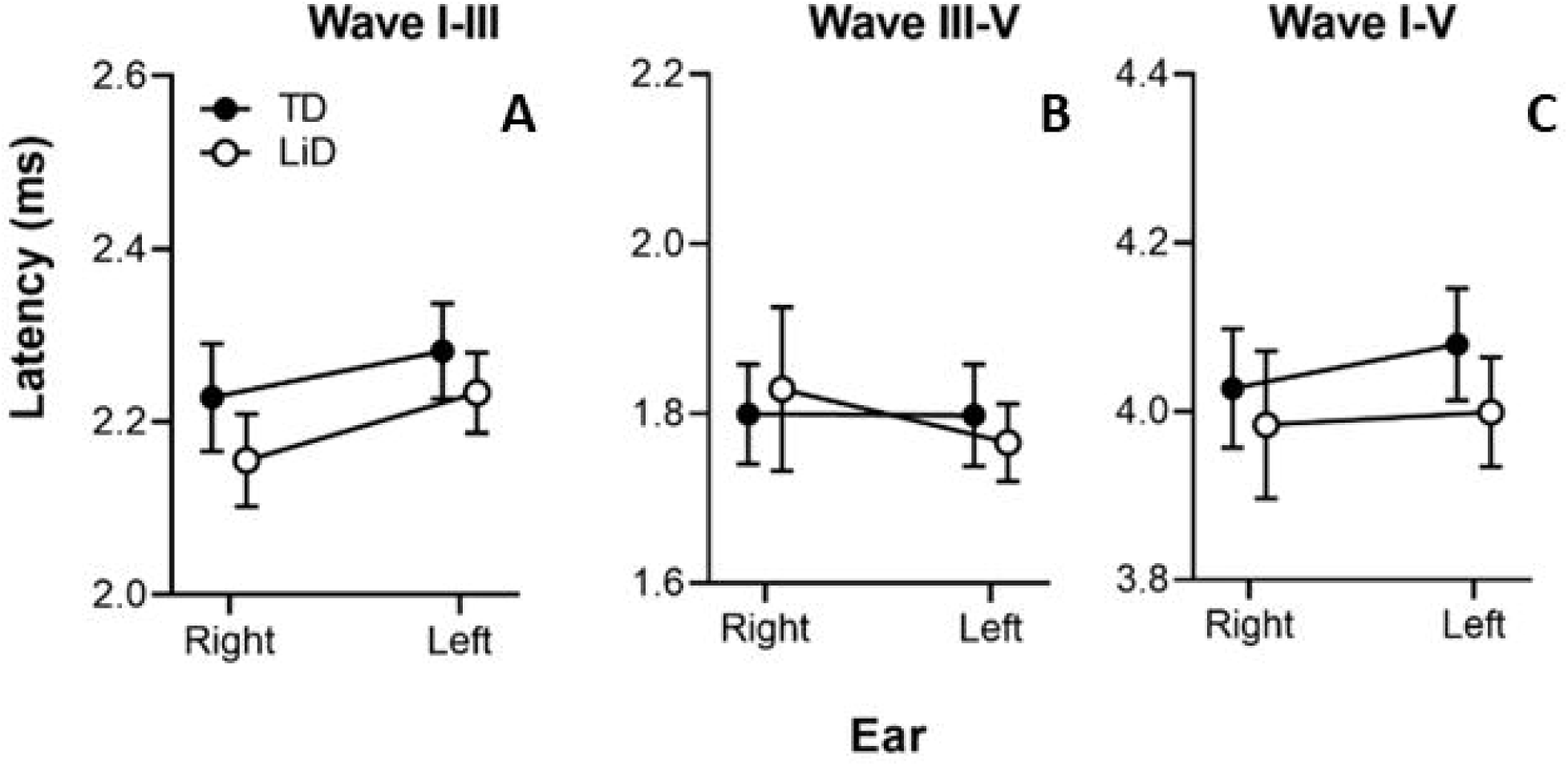
Ipsilateral ABR interpeak latencies for Waves I, III and V for ear. The LiD and TD group are shown in open and filled circles, respectively. Error bars are 95% confidence intervals. N= 67 ears for the TD group and 66 ears for the LiD group. Overall ANOVA: Group p=0.089, Ear p=0.357.

**Figure 4.**
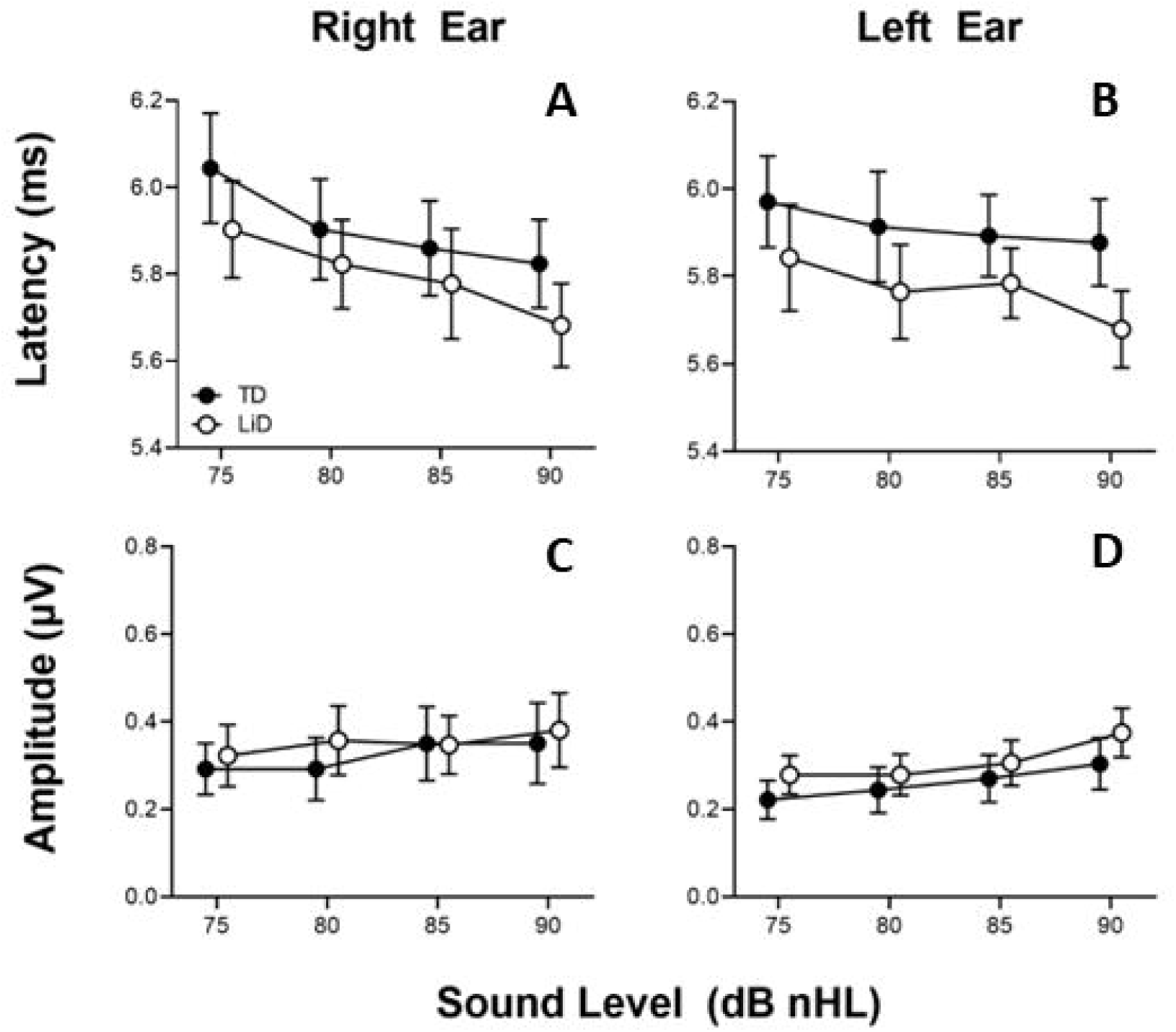
Contralateral ABR latency-intensity and amplitude-intensity functions for Wave V in dB HL re: normal adult threshold for clicks. The LiD and TD group are shown in open and filled circles, respectively. Error bars are 95% confidence intervals. N= 67 ears for the TD group and 66 ears for the LiD group.

**Table 2.** ABR repeated measures analysis of variance *p-* and F-values for factors included in the final model.

|  | Intensity | Group | Ear | Sex | Tube History | EHF HL |
| --- | --- | --- | --- | --- | --- | --- |
| **Ipsilateral** |  |  |  |  |  |  |
| SP Latency (p) | < .001 | 0.436 | **0.017** | 0.934 | 0.722 | 0.216 |
| F (DF = 1 to 3.0) | **69.262** | 0.613 | **5.932** | 0.007 | 0.127 | 1.552 |
| SP Amplitude (p) | < .001 | 0.532 | 0.093 | 0.086 | 0.110 | 0.321 |
| F (DF = 1 to 3.0) | **103.104** | 0.393 | 2.886 | 3.011 | 2.599 | 0.998 |
| Wave I Latency (p) | < .001 | 0.635 | 0.046 | 0.982 | 0.943 | 0.124 |
| F (DF = 1 to 2.5) | **136.889** | 0.226 | 4.061 | < .001 | 0.005 | 2.403 |
| Wave III Latency (p) | < .001 | **0.001** | 0.582 | 0.228 | 0.139 | 0.400 |
| F (DF = 1 to 2.5) | **62.270** | **10.809** | 0.304 | 1.472 | 2.219 | 0.715 |
| Wave V Latency (p) | < .001 | **0.036** | 0.850 | **0.030** | 0.745 | 0.175 |
| F (DF = 1 to 2.3) | **60.952** | **4.497** | 0.036 | **4.818** | 0.107 | 1.865 |
| Wave I Amplitude (p) | < .001 | 0.317 | 0.132 | **0.002** | 0.089 | 0.349 |
| F (DF = 1 to 2.6) | **76.639** | 1.012 | 2.297 | **9.906** | 2.945 | 0.885 |
| Wave III Amplitude (p) | **0.002** | 0.969 | CNT | 0.372 | 0.173 | 0.173 |
| F (DF = 1 to 3.0) | **5.762** | 0.002 | CNT | 0.837 | 2.002 | 2.006 |
| Wave V Amplitude (p) | < .001 | 0.280 | 0.297 | **0.010** | 0.438 | 0.421 |
| F (DF = 1 to 2.6) | **21.270** | 1.178 | 1.098 | **6.794** | 0.605 | 0.652 |
| Wave V/I Amplitude (p) | 0.918 | 0.735 | 0.107 | 0.701 | 0.914 | 0.850 |
| F (DF = 1 to 2.6) | 0.168 | 0.115 | 2.643 | 0.148 | 0.012 | 0.036 |
| Interpeak Latency (p) | NA | 0.089 | 0.357 | **0.029** | 0.673 | 0.479 |
| F (DF = 1 to 1.3) | NA | 2.933 | 0.857 | **4.870** | 0.179 | 0.504 |
| **Contralateral** |  |  |  |  |  |  |
| Wave V Latency (p) | < .001 | 0.093 | 0.877 | 0.126 | 0.316 | **0.031** |
| F (DF = 1 to 2.4) | **20.949** | 2.890 | 0.024 | 2.390 | 1.019 | **4.780** |
| Wave V Amplitude (p) | < .001 | 0.457 | 0.038 | 0.707 | 0.241 | 0.444 |
| F (DF = 1 to 3.0) | **9.637** | 0.563 | 4.531 | 0.143 | 1.409 | 0.596 |

*Note:* CNT = Could not test; DF = Degrees of Freedom; EHFHL = Extended High Frequency Hearing Loss (> 20 dB HL, 10 to 16 kHz); Bold italics indicate significant *p*-values.

Due to the significant latency group differences for ipsilateral ABR Waves III and V (RMANOVA), correlations with the Total ECLiPS score across all participants were examined using multiple forward stepwise regression. Shorter Wave V latency was correlated significantly with lower ECLiPS Total scores for the right ear (Figure 5A; r = 0.296, *p* = 0.023). Correlations for right ear Wave III and for left ear Waves III and V latency did not reach significance. Correlations between ABR Wave III or V with subtests of the LiSN-S test (low cue, high cue, talker, and spatial advantage scores) were not significant for either ear.

**Figure 5.**
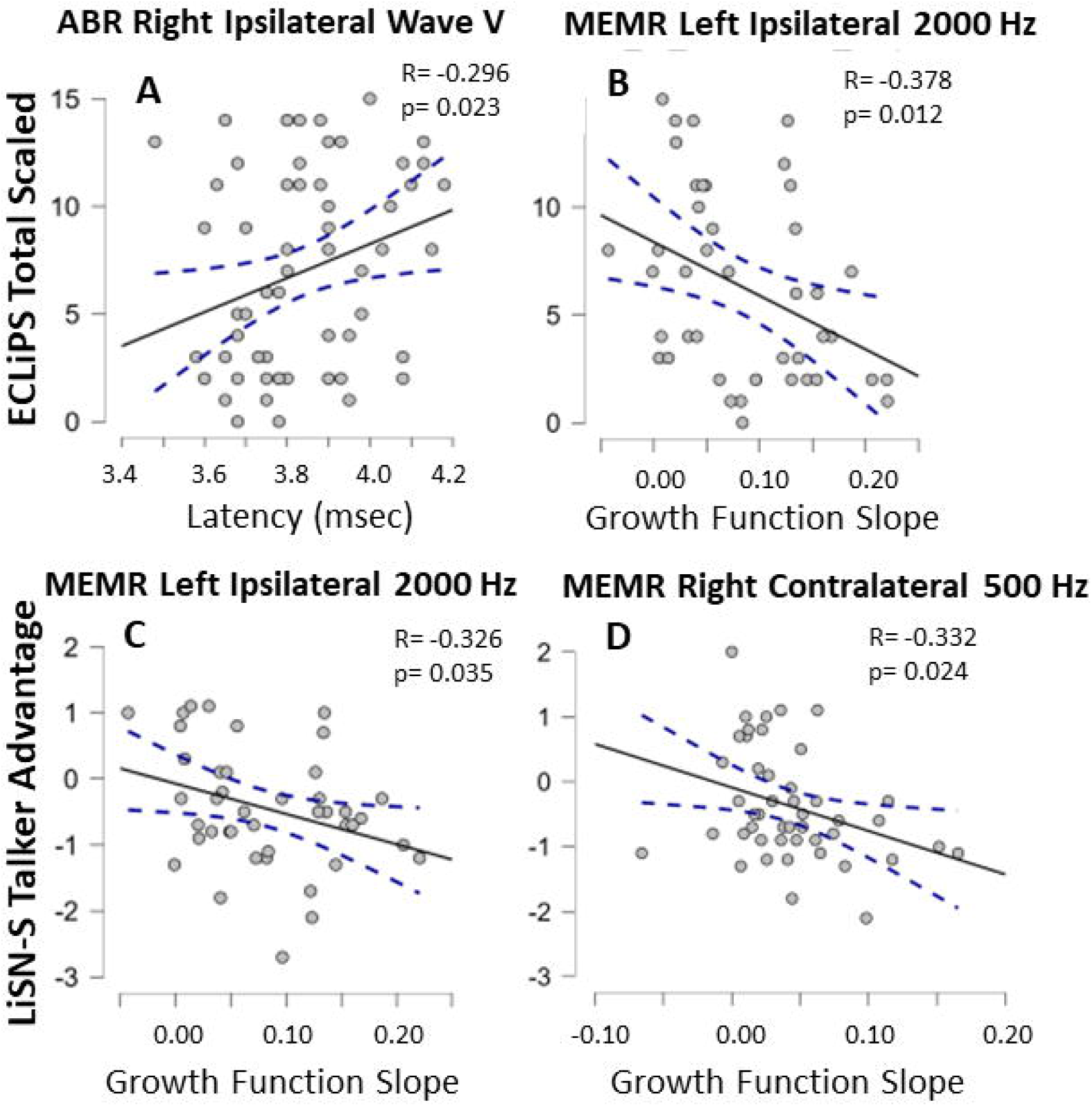
Correlations between ABR wave V and the ECLiPS scale (A); between MEMR left ipsilateral growth function slope and the ECLiPS (B); between left ipsilateral growth function slope and the LiSN-S talker advantage (C); and between right contralateral growth function slope and the LiSN-S talker advantage (D).

### 3.3 Middle Ear Muscle Reflex

MEMR responses were obtained from 54 LiD participants (n = 106 ears) and 49 TD participants (n = 95 ears). Ipsilateral and contralateral MEMR slope functions (steepness of amplitude growth as the stimulus is increased) were examined with separate RMANOVAs. Stimulus type (BBN, 0.5, 1, and 2 kHz) was the repeated factor, group and ear were between subject factors, while age, sex, average EHF hearing levels (10-16 kHz) and tube history were included as covariates. Mean MEMR slope values, separated by laterality, stimulus type, and test ear, are shown in Figure 6 with the RMANOVA results in Table 3.

**Figure 6.**
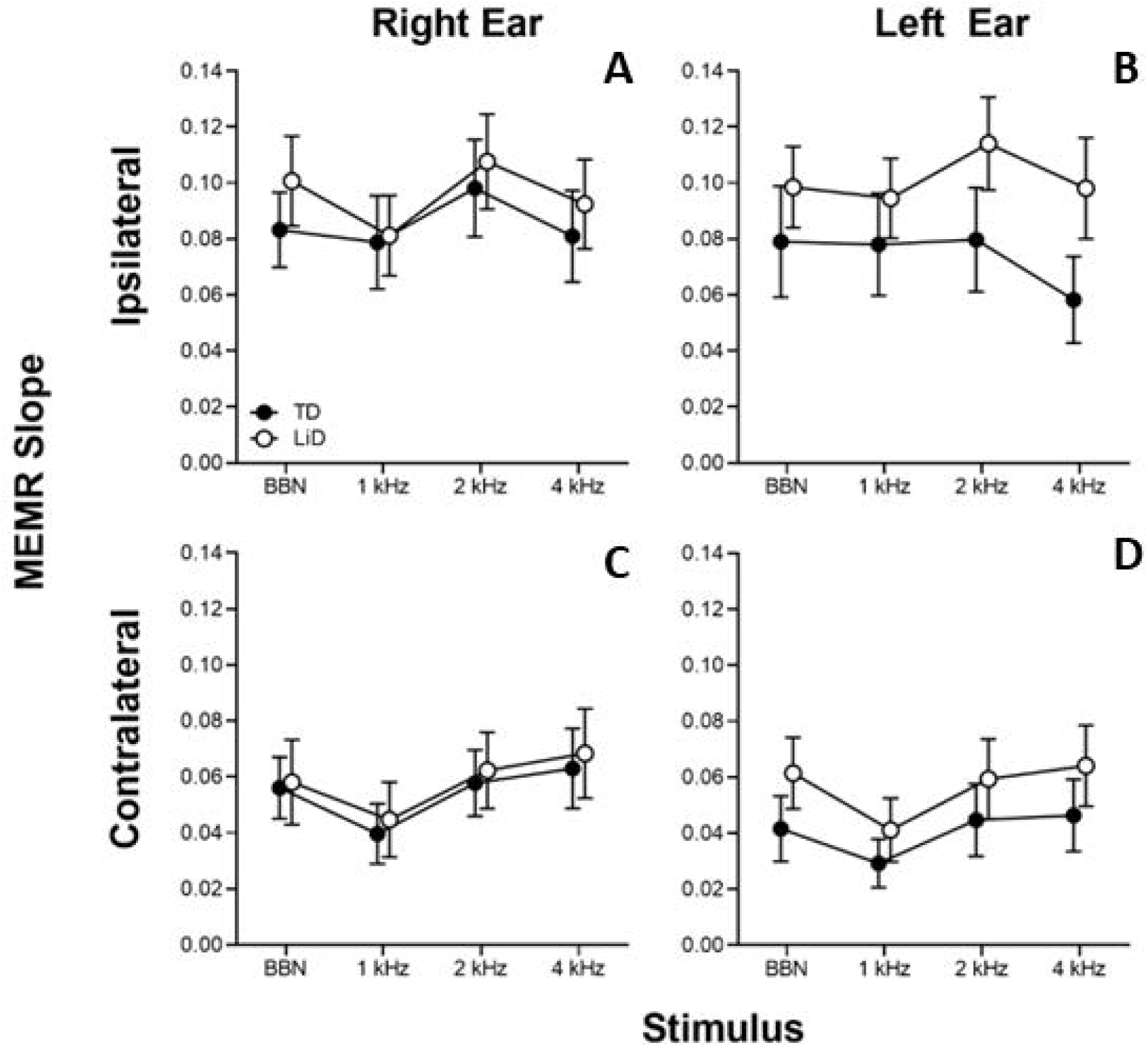
Wideband middle ear muscle reflex (MEMR) slope functions for ipsilateral (A, B) and contralateral (C, D) recording modes. The LiD and TD group are shown in open and filled circles, respectively. Error bars are 95% confidence intervals. N= 79 ears for the TD group and 87 ears for the LiD group.

**Table 3.** MEMR growth slope repeated measures analysis of variance, with *p-*values and F-test (Degrees of Freedom) displayed for factors included in the final model.

|  | Stimulus<br>Type | Group | Ear | Sex | Tube<br>History | EHF HL | Age |
| --- | --- | --- | --- | --- | --- | --- | --- |
| **Ipsilateral** |  |  |  |  |  |  |  |
| MEMR Slope (p) | ***0.017*** | ***0.035*** | 0.533 | ***0.014*** | ***0.015*** | 0.205 | 0.245 |
| F (DF = 1 to 2.6) | ***3.631*** | ***4.537*** | 0.389 | ***6.134*** | ***6.054*** | 1.618 | 1.362 |
| **Contralateral** |  |  |  |  |  |  |  |
| MEMR Slope (p) | 0.448 | ***0.025*** | ***0.026*** | ***0.009*** | ***0.049*** | ***< .001*** | 0.610 |
| F (DF = 1 to 2.7) | 0.867 | ***5.149*** | ***5.066*** | ***7.032*** | ***3.939*** | ***21.026*** | 0.261 |

*Note:* Stimulus Condition = BBN, 0.5, 1, 2 kHz; EHFHL = Extended High Frequency Hearing Loss (> 20 dB HL 10 to 16 kHz) from; Bold italics indicate significant p-values.

Significant effects of stimulus type, group, sex, and PE tube history were found (Figure 6; A, B). LiD participants showed steeper MEMR slope, averaged across frequency (Mean = 0.10; SD = 0.05) compared to TD participants (Mean = 0.08; SD = 0.05). Females had a steeper MEMR slope (Table 3; Mean = 0.10; SD = 0.06) compared to males (Mean = 0.08 SD = 0.07). Conversely, individuals with a history of tubes had a shallower MEMR slope (Mean = 0.06; SD = 0.04) compared to individuals with no tube history (Mean = 0.10, SD = 0.05). A significant interaction was found between group and ear, specifically, participants with LiD had steeper MEMR slope in the left ear (Mean = 0.10, SD = 0.05) compared to TD participants (Mean = 0.07, SD = 0.05, *p* = 0.023). No other group by ear interactions were significant. Post-hoc analysis showed that the 1kHz stimulus (Mean = 0.10; SD = 0.06) had greater slope compared to 2kHz (Mean = 0.08; SD = 0.06; *p* = 0.004), .5 kHz (Mean = 0.08; SD = 0.05; *p* = 0.004) and BBN (Mean = 0.09, SD = 0.05; *p* = 0.043).

For contralateral MEMR slope, there was no significant effect of stimulus type (Figure 6 C, D: Table 3). However, there were significant effects of group, ear, gender, tube history and average EHF hearing level. Participants with LiD showed significantly steeper MEMR slopes (Mean = 0.06; SD = 0.04) compared to TD participants (Mean = 0.05; SD = 0.03). The MEMR slope was steeper in the right ear (Mean = 0.06; SD = 0.040) compared to the left ear (Mean = 0.04; SD = 0.04). Females had steeper MEMR slope (Mean = 0.06; SD = 0.04) compared to males (Mean = 0.04; SD = 0.03). Conversely, individuals with a history of tubes had shallower MEMR slopes (Mean = 0.03; SD = 0.03) compared to individuals with no tube history (Mean = 0.06, SD = 0.04). Similarly, individuals with poorer EHF hearing had shallower MEMR growth compared to individuals with better EHF hearing (r = -0.361; *p* < 0.001).

Correlations between MEMR and other measures were examined across all participants using multiple forward stepwise regression for each “family” of tests to control for covariance (multiple stimulus frequencies within each MEMR test) and the ECLiPS standard score, and for each subtest of the LiSN-S test. There was a significant correlation between the ipsilateral left ear 2 kHz MEMR slope and ECLiPS scores, shown in Figure 5 B. Correlations were also examined for all LiSN-S subtest scores and MEMR. Significant correlations were found between ipsilateral left ear 2 kHz MEMR slope and LiSN-S talker advantage scores (Figure 5 C; r= - 0.326, p= 0.035). There was also a significant correlation between contralateral right ear MEMR growth at 0.5 kHz and LiSN-S test talker advantage (r= -0.332, p= 0.024). All other correlations were not significant.

### 3.4 Binaural Medial Olivocochlear Reflex

Binaural MOCR values are displayed as dB of suppression compared to the baseline response (Figure 7). Independent sample t-tests were conducted for the left and right ear separately to examine group differences. Results showed no significant MOCR group differences in the left (*p* = 0.636) or right ear (*p* = 0.314).

**Figure 7.**
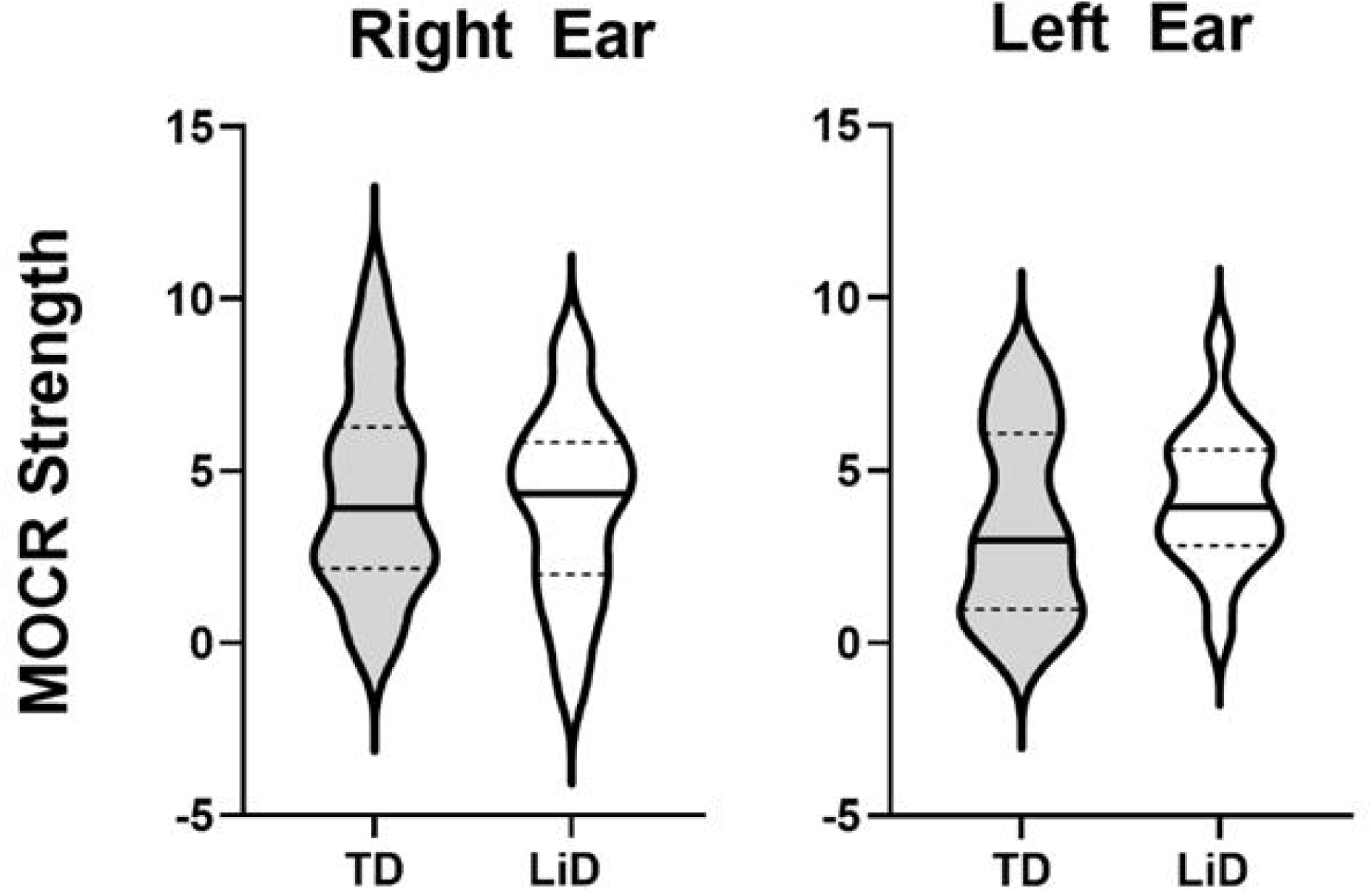
Violin plots for the binaural medial olivocochlear reflex (MOCR) expressed in dB of suppression compared to baseline response. The plots show median center line, interquartile range (box), 95% range (stems), and distribution (outlines). Results for the left ear based on n=22 ears for the TD group and 17 ears for the LiD group (p=.314). Results for the right ear based on n=26 ears for the TD group and 22 ears for the LiD group (p=.636).

### 3.5 Envelope Following Response

EFR amplitude and phase locking value (PLV) as well as the EFR and PLV signal to noise ratio (SNR) are displayed as a function of modulation depth in Figure 8, and the RMANOVA results are displayed in Table 4. Separate RMANOVAs were conducted for EFR amplitude, EFR SNR, PLV, PLV SNR with modulation depth (100%, 63%, and 40%) as the repeated condition. Group was the between subject factor, and age, sex, average EHF hearing levels (10-16 kHz) and tube history were covariates.

**Figure 8.**
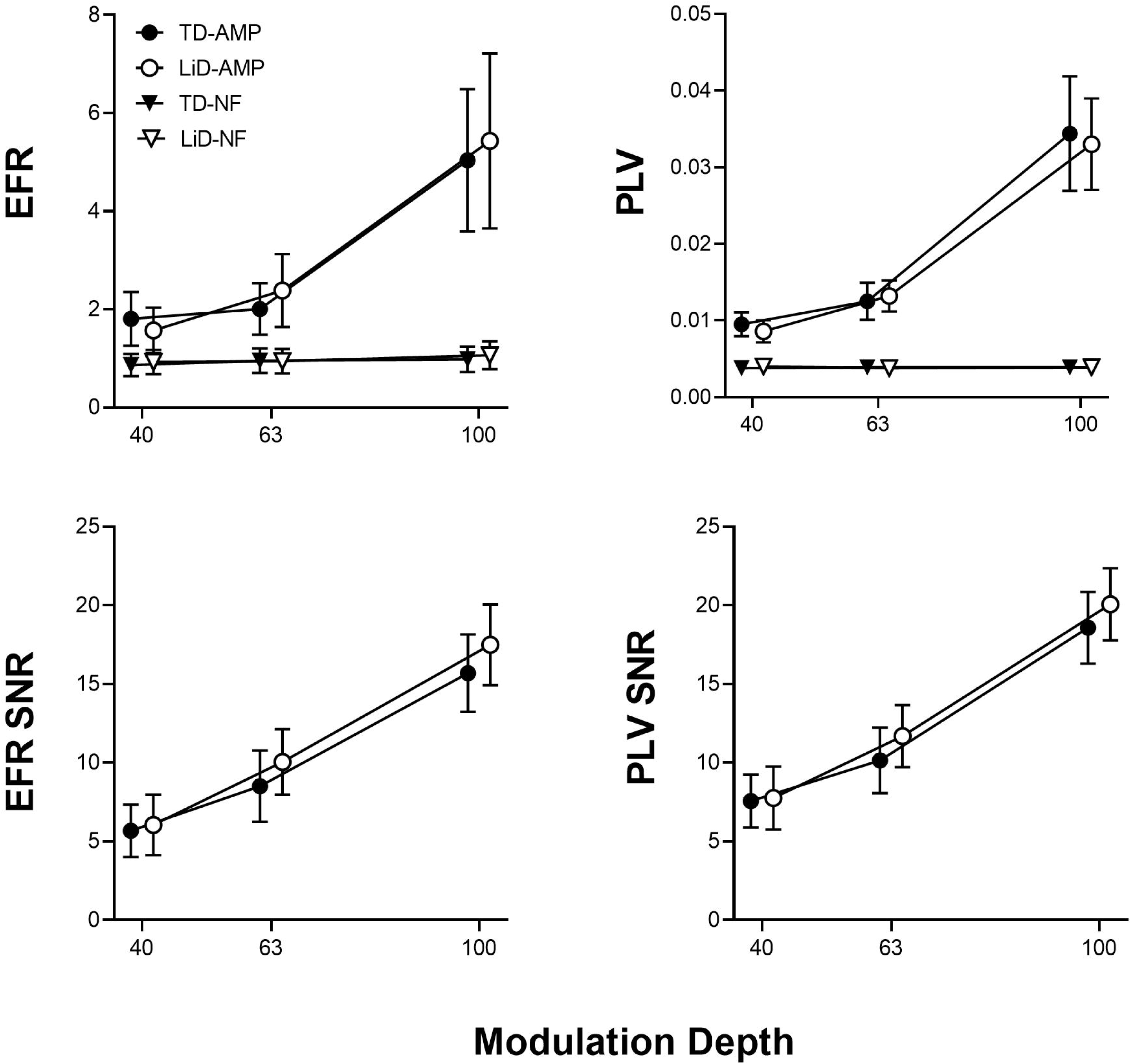
Top panels: Envelope following response (EFR) as a function of modulation depth for phase locking value (PLV; A) and EFR amplitude (B) by group, N= 25 ears (TD); 30 ears (LiD). Overall phase locking value analyzed by group p=.319. Overall EFR amplitude analyzed by group p=.434. EFR signal to noise ratio (SNR) as a function of modulation depth for phase locking value (PLV; C) and EFR amplitude (D) by group, N= 25 ears (TD); 30 ears (LiD). Overall phase locking value analyzed by group p=0.851. Overall EFR amplitude analyzed by group p=0.186.

**Table 4.** EFR repeated measures analysis of variance, with *p-*values and F-test (Degrees of Freedom) displayed for factors included in the final model.

|  | Modulation<br>Depth | Group | Sex | Tube History | EHFHL | Age |
| --- | --- | --- | --- | --- | --- | --- |
| **EFR** |  |  |  |  |  |  |
| Amplitude (p) | ***0.002*** | 0.186 | 0.221 | 0.257 | 0.359 | 0.057 |
| F (DF = 1 to 1.1) | ***9.843*** | 1.812 | 1.544 | 1.326 | 0.863 | 3.834 |
| SNR (p) | ***0.021*** | 0.482 | 0.317 | 0.862 | 0.385 | 0.475 |
| F (DF = 1 to 1.6) | ***4.474*** | 0.503 | 1.024 | 0.031 | 0.771 | 0.520 |
| **Phase Locking** |  |  |  |  |  |  |
| Value (p) | 0.179 | 0.851 | 0.909 | 0.373 | 0.270 | 0.867 |
| F (DF = 1 to 1.1) | 1.868 | 0.036 | 0.013 | 0.810 | 1.251 | 0.028 |
| SNR (p) | ***0.019*** | 0.669 | 0.821 | 0.521 | 0.513 | 0.548 |
| F (DF = 1 to 1.7) | ***4.526*** | 0.185 | 0.052 | 0.419 | 0.434 | 0.367 |

*Note:* EHFHL = Extended High Frequency Hearing Loss (> 20 dB HL, 10 – 16 kHz); Bold italics indicate significant *p*-values.

No significant main group differences were found for EFR amplitude, PLV amplitude, EFR SNR and PLV SNR. There were no significant effects of sex, tube history, average EHF hearing level or age at test for the EFR or PLV measures. As expected, modulation depth EFR amplitude and SNR significantly increased with modulation depth, as did PLV SNR.

## 4.0 DISCUSSION

Shorter ABR Wave III and V latency and steeper MEMR growth were found in the LiD group; these enhanced responses were related to poorer ECLiPS scores and poorer talker advantage on the speech in noise task. The finding of shorter ABR wave III and V latency for the LiD group was surprising, and in the opposite direction to that hypothesized, as delay in the ascending auditory pathway would logically relate to listening difficulties, with longer latencies reflecting slower speed of neuronal transmission. However, a previous study in 10 normal hearing children with learning disabilities and suspected APD compared to 10 age and gender matched control children also reported significantly shorter Wave V latency and Wave III-V latency in the APD group (Purdy, Kelly, & Davies, 2002). The significant correlation between parent report of LiD and ABR Wave V latency, and between speech in noise and MEMR growth has not been previously reported.

In contrast, previous studies of ABR latency in children with APD have mostly found normal results. In a study of children with suspected APD, and with or without a defined diagnosis, ABR latencies and amplitudes were not related to diagnosis (Allen & Allan, 2014). A study of 20 children diagnosed with APD, tested with electrocochleography and ABR, found no significant absolute latency or interpeak latency group differences, compared to 16 typically developing children and 20 normal hearing adults (Veeranna, Allan, & Allen, 2021). Significantly smaller Wave V amplitudes were reported in the APD cases, but there was not a significant Wave V/I amplitude difference. A retrospective study of 108 children suspected of APD and tested with click ABR showed no significant group difference from typically developing children and adults, but some individual children (37%) were reported to have delayed latencies and less stable responses (Ankmnal-Veeranna et al., 2019). Similarly, a study of 19 children diagnosed with APD found no significant ABR latency or amplitude differences compared to 24 controls (Morlet et al., 2019).

In a similar vein, if neural evidence for LiD were present in the lower brainstem, MEMR growth with stimulus level would be predicted to be shallower in children with LiD. However, we found the opposite – significantly steeper MEMR growth with increasing level in the LiD group for both ipsilateral and contralateral MEMR. In contrast, shallower contralateral MEMR growth curves were reported in children with suspected APD (Saxena et al., 2015). Interestingly, we found significantly steeper MEMR growth slopes in the left ear, compared to the right ear, but only in the LiD group. Of further interest is the relationship between poorer ECLiPS scores and steeper ipsilateral left ear MEMR, and between poorer talker advantage on the LiSN-S and steeper contralateral right and ipsilateral left ear MEMR responses. Thus, there was a complex mix of significant relations between behavioral measures and MEMR growth. This was a clear divergence from peripheral effects, in that poorer EHF hearing was associated with shallower MEMR responses, and PE tube history related to shallower MEMR responses. This divergent relationship between peripheral hearing loss and MEMR growth curves suggests a central origin for the finding of steeper growth in children with LiD, reflecting decreased efferent control or, alternatively, increased central gain.

EFR and PLV measures have been proposed as sensitive to cochlear synaptopathy (Bharadwaj et al., 2014; Shaheen et al., 2015). We therefore included those measures in this study on the premise that CS may contribute to LiD. However, we found no evidence for the impaired temporal encoding that PLV is sensitive to in the LiD group.

In terms of peripheral hearing effects, two measures were related to EHF hearing levels for both groups combined – longer ABR contralateral Wave V latency, and shallower MEMR growth and slope. Thus, some children in the LiD and TD groups showed a significant effect of poorer EHF hearing thresholds, despite having normal standard frequency thresholds. Poorer EHF hearing was related to longer ABR latency and shallower MEMR growth, a novel finding that may indicate diffuse cochlear deficits despite normal hearing in the standard frequency range, as the stimuli used for ABR and MEMR are primarily below 4 kHz. Evidence has accumulated from recent reports that EHF hearing is related to speech in noise SRT (Flaherty, Libert, & Monson, 2021; Motlagh Zadeh et al., 2019; Polspoel, Kramer, van Dijk, & Smits, 2021) and to a range of physiologic measures in the standard frequency range. EHF is also an important early marker of peripheral auditory damage (Blankenship et al., 2021; Hunter, Monson, et al., 2020; Mishra, Saxena, & Rodrigo, 2022; Motlagh Zadeh et al., 2019).

The binaural MOCR paradigm we used has been previously shown to produce the largest and most reliable effect of the different MOCR methods (Lilaonitkul & Guinan, 2009). Despite rigorous quality control measures for attention, SNR, sufficient averaging, and control of the MEMR threshold, only 38% of MOCR recordings overall met quality criteria to be included in the analysis. This is a significant limitation of these results, so interpretation must be cautious with respect to an absence of group significance. Test-retest reliability was poorer in the LiD group and may be related to poorer attention (Moore et al., 2010; Stavrinos, Iliadou, Edwards, Sirimanna, & Bamiou, 2018) and greater internal noise (Porter, Leibold, & Buss, 2020) in that group. It may also be the case that the MOCR as measured by the TEOAE suppression method is not related to difficulties in natural listening situations, consistent with some previous studies (Mattsson et al., 2019; Smart, Kuruvilla-Mathew, Kelly, & Purdy, 2019).

A recent investigation by Rao, Koerner, Madsen, and Zhang (2020) in adults reported no significant effect of contralateral MOCR activation on auditory cortical responses or on a nonsense word recognition in noise task. They concluded that the MOCR may not play a primary role in higher level processing of speech in noise perception. Related to this hypothesis, Hernandez-Perez et al. (2021) measured MOCR strength using TEOAE suppression and compared it to neural activity along the ascending pathways in response to “degraded” (vocoded) or conventional noise-masked speech. The MOCR was activated by the vocoded speech signal, but not by speech-in-noise which, instead, increased neural activity in the midbrain and cortex. They suggested that the auditory system has distinct strategies to handle these two types of distorted speech. In the ear and hindbrain, the MOCR may enhance the stimulus waveform, whereas the mid- and forebrain may be specialized to reduce the effect of added noise.

Our finding of no significant differences between groups for lower-level auditory mechanisms (MOCR, EFR, SP and Wave I) is consistent with the hypothesis that lower-level auditory pathway function does not contribute to the LiD problems experienced by children in this study. Group differences emerged, however, as we ascended the auditory brainstem to levels involved in ABR Waves III and V, and MEMR growth. Poorer subcortical control and left-right balance of auditory information was indexed by 1) shorter Wave III and V latencies, with right Wave V related to poorer caregiver reports of LiD, and 2) steeper MEMR growth, related to performance in a natural, sentence-in-distractor task and the validated parent reports of LiD. These findings suggest the possibility that children with LiD have atypical corticofugal mechanisms. Poorer corticofugal control is consistent with a range of other evidence for poorer cortical function in children with LiD (Dawes & Bishop, 2009; Ferguson, Hall, Riley, & Moore, 2011; Miller & Wagstaff, 2011; Moore, Ferguson, Edmondson-Jones, Ratib, & Riley, 2010; Petley et al., 2021b; Sharma, Purdy, & Kelly, 2009).

As reviewed by Souffi et al. (2021), recent animal evidence suggests that, during active listening, the frontal cortex activates the auditory cortex (AC) and IC directly and, via the AC, the MGB and IC indirectly, producing strong top-down control. Because the ABR and MEMR measures we employed here did not involve active listening to natural speech in noise, it seems unlikely that such downward control mechanisms were operational. Rather, the shorter latency and steeper growth curves in the LiD group may reflect poorer function of the corticofugal suppressive control networks (Souffi et al., 2021) having further downstream influences on brainstem ABR Waves III and V, and the MEMR. We are currently in the process of analyzing active speech-evoked and passive resting state MRI in these same children and will evaluate this hypothesis regarding functional connectivity of the frontal regions, primary auditory cortex, and auditory thalamus.

## 5.0 CONCLUSIONS

In conclusion, evidence was found for mid-brainstem enhancements of ABR Wave III and V and MEMR, suggesting corticofugal differences between children with and without LiD. These differences related, in turn, to parent report of listening problems and poorer perception of natural speech in competing masker speech. These results suggest that MEMR growth curves may serve as a metric of efferent activity. The fact that they also relate to behavioral function is an important, novel finding. These findings have implications for treatments focused on improving speech in noise performance, ideally during earlier childhood when neuroplasticity can be maximized.

## Data Availability

All data produced in the present study are available upon reasonable request to the authors.

## Acknowledgments

Supported by NIH R01DC014078 (Moore), NIH 2UL1TR001425-05 (University of Cincinnati), and the Cincinnati Children’s Research Foundation. The views expressed do not necessarily represent the National Institutes of Health. Portions of this study were presented as poster presentations at the Association for Research in Otolaryngology (2017; 2018; 2021) and American Auditory Society (2018). Thanks to Douglas Keefe for providing the wideband immittance software and consultation. Nicholette Sloat, MS and Audrey Perdew, MS, from the Moore lab, Morgan Bamberger, BS from the Hunter Lab and Laura Tatro, AuD from the Hood Lab assisted with data collection and analyses. David Moore receives support from the NIHR Manchester Biomedical Research Centre. We also thank our participating families and UC as well as Summer Undergraduate Research Foundation (SURF) scholars.

## Author Contributions

David Moore and Lisa Hunter: Conceptualization; Data curation; Formal analysis; Investigation; Methodology; Project administration; Resources; Supervision; Validation; Writing original draft; review & editing. Linda Hood and Barbara Shinn-Cunningham designed experiments and analysis procedures. Chelsea Blankenship collected data, analyzed data and writing, editing. Lina Motlagh Zadeh: Data collection and editing.

## Open Access

Data are available upon reasonable request to the corresponding author.

## Notes

**Conflicts of Interest and Source of Funding:** This research was supported by the National Institute of Deafness and other Communication Disorders of the National Institutes of Health under Award Number R01 DC014078 and the Cincinnati Children’s Hospital Medical Center Research Foundation. There are no conflicts of interest to disclose.

### Competing Interest Statement

The authors have declared no competing interest.

### Funding Statement

This research was supported by the National Institute of Deafness and other Communication
Disorders of the National Institutes of Health under Award Number R01 DC014078 and the National Center for Advancing Translational Sciences of the National Institutes of Health, under Award Number UL1TR001425. The content is solely the responsibility of the authors and does not necessarily represent the official views of the NIH.

### Author Declarations

Ethics committee/IRB of Cincinnati Children's Hospital Medical Center gave ethical approval for this work.

